# Global health security capacity against COVID-19 outbreak and its association with the case fatality rate: an analysis of annual data from 210 countries and territories

**DOI:** 10.1101/2020.04.25.20079186

**Authors:** Elham maraghi, Amal Saki Malehi, Fakher Rahim

## Abstract

**Background:** Because infectious diseases, such as COVID-19, do not have specific boundaries, all countries must prioritize and use the necessary capabilities to prevent, detect, and respond quickly to public health emergencies. In this context, we aimed to review most recent GHS index annual report to observe the regional and global level of health security against COVID-19 outbreak, as well as their relationship with case fatality rate, among 210 countries and territories worldwide.

**Methods:** We reviewed and analyzed October 2019 GHS index co-leaders joint report, to review health security capacities on the basis of the GHS index in the context of six categories. we prioritized not only the capacities of 210 countries and territories around the world using the GHS Index, but also the existence of functional, tested, proven capabilities for stopping outbreaks at the source. Data were collected from global databases including Worldometer, WHO, and Disease Control and Prevention Center (CDC).

**Findings:** This study recruited data on 210 countries and territories, of which up to 14 April 2020, 72 countries (34.28%) with more than 1000 total COVID-19 cases were presents. In “most prepared group”, number of total COVID-19 diagnostic tests had a significant positive relation with GHS index (r=0.713; p=0.006). Case fatality rate was directly associated with the detection index (r=0.304; p=0.023) in “more prepared group”. In “Lower-middle-income economies” group, case fatality rate positively related to detection, response and risk environment indices.

**Implementation:** With the exception of a very small number, countries that were ranked as most prepared countries, they were more likely to be affected by the COVID-19 outbreak of the virus and its health consequences, and needed to seriously reconsider their capabilities and health security in the context of detection, prevention, rapid response, health system facilities, and risk environment against disease outbreak

**Research in context:** *Evidence before this study:* Given the very rapid spread of the COVID-19 disease in a very short time, limited and few studies have shown weakness and strength in national and international capacity to deal with health emergencies. We systematically searched the Scopus, ISI web of science and PubMed from Jan 2019 to April 2020, using the search terms “health security” OR “emergency preparedness” AND “COVID-19” OR “SARS-CoV-2/nCoV-2019”. Our search returned only limited number of published evidences (n=37), of which only one was assessed the operational readiness among 182 countries based on the International Health Regulations (IHR) annual report ^1^.

*Added value of this study:* Given a very limited and insufficient on the regional, as well as global preparedness capacities to combat health emergencies, such as COVID-19 disease, we used most recent GHS index annual report (October 2019), to observe the regional and global level of health security in the context of detection, prevention, rapid response, health system facilities, and risk environment against COVID-19 outbreak among 210 countries and territories around the world. We found information about only 195 countries in the recent used report and imputed the data for the rest 15 countries and territories that facing COVID-19 outbreak.

*Implications of all the available evidence:* Our results showed that, with the exception of a very small number of countries that were ranked as most prepared countries, they were more likely to be affected by the COVID-19 outbreak of the virus and its health consequences, and needed to seriously reconsider their capabilities and health security in the context of detection, prevention, rapid response, health system facilities, and risk environment against disease outbreak.

## Introduction

The World Health Organization (WHO) announced on March 11, 2020 that the outbreak of the 2019 novel coronavirus (COVID-19) has turned to a global epidemic ^2^. In the last two weeks, the number of people infected with the virus has tripled and the number of countries affected has reached 210. To date, more than 1.8 million cases have been reported in 210 countries, of which about 1,400,000 have died ^3^. The symptoms of the virus are 80% similar to the acute respiratory syndrome of SARS, and the virus transmits through coughing and sneezing ^4^. The symptoms of the early stages of COVID-19 infection occurs in the form of high fever, sore throat, shortness of breath and diarrhea. In the later stages, it causes pneumonia and kidney failure, and could eventually lead to the patient’s death ^5^. There is currently no vaccine or effective treatment for the virus, and the only way to prevent is to diagnose the patient early and quarantine the confirmed as well as suspected cases ^6^. Authorities in some countries have asked citizens to refrain from attending crowded and public places. The virus has also spread to several countries through travelers, raising concerns around the world and treated the global health security.

Because infectious diseases, such as COVID-19, do not have specific boundaries, all countries must prioritize and use the necessary capabilities to prevent, detect, and respond quickly to public health emergencies. Each country must also be transparent about its capabilities to reassure its neighbors that it can prevent the spread of an international catastrophe. In turn, global leaders and international organizations have a collective responsibility to develop and maintain a strong global ability to counter the threat of infectious diseases. This ability includes ensuring that financial resources are provided to fill the gap in the readiness for epidemics, which saves human lives and achieves a safer and more secure world. The Global Health Security (GHS) index is the first comprehensive indicator and benchmark for assessing health security and related capabilities in 195 countries; therefore, this index is a necessary tool to deal with the risks of transnational public health, of which uses to prevent, protect, control and respond without disrupting international trade. In this index, biological events with high consequences are defined as the prevalence of infectious diseases that can overshadow the national or international capacity to manage them.

The unique feature of the GHS index is that it provides a comprehensive assessment of countries’ health security and considers the broader context of biological hazards in each country, including a country’s geopolitical considerations and health-care system. However, knowing the dangers is not only not enough, but political will is needed to protect the people from the consequences of epidemics as well as pandemics, to save lives and to create a safer and more secure world. Delays in the global response to the COVID-19 pandemic could lead to a shift in the structure of the world and call for a transparent assessment of countries’ public health capacities. So far, health-related, policy and security agencies have provided a number of high-level studies, and have identified ways to identify, fund, and fill key gaps in preparedness. These recommendations for epidemic threats, such as COVID-19, could have a geographical scope, intensity, or social impact, as well as overshadow national or international capacity to manage it. The recommendations have been provided, but many of them have not yet been implemented due to lack of funding.

Given the importance of the GHS index as a tool to fully assess the health systems of different countries and their capacities to control the spread of the disease, the existing capacities and programs of countries and their readiness to understand the comprehensive effects of epidemics, their specific measures provide credibility and fill in the gaps in the anti-epidemic system, indicators and questions that form the framework of this index, as well as analyze the capacity of health security within the context of a national health systems and other risk factors, could be a global priority. Therefore, this study aims to calculate and evaluate the GHS index in countries affected by the COVID-19 outbreak and its public health related consequences, as well as its relationship with the case fatality and recovery rates using up-to-date data at the global.

## Methods

We reviewed and analyzed October 2019 GHS index co-leaders joint report in contribution with Nuclear Threat Initiative (NTI) and Center for Health Security (CHS), Johns Hopkins Bloomberg School of Public Health, to review health security capacities on the basis of the GHS index in the context of six categories, including prevention (preventing the emergence or release of pathogens), early detection and reporting (early detection and reporting of epidemics and potential international concern), rapid response (timely response and reduction of the spread of a pandemic disease), health system (sufficient and strong health system for the treatment of patients and protection of health workers), compliance with international standards and norms (commitments to improve national capacity, funding programs to bridge the gap, and compliance with global norms), and the risk environment (general vulnerability of the environment and vulnerability of the country to biological threats), is reviewed and summarized. Among its 140 questions, we prioritized not only the capacities of 210 countries and territories around the world using the GHS Index, but also the existence of functional, tested, proven capabilities for stopping outbreaks at the source. The overall GHS Index, as well as prevention, detection and reporting, rapid response, health system, compliance with international standards and norms, and the risk environment scores among all 195 countries were considered. The average overall GHS Index, prevention, early detection and reporting, rapid response, health system, compliance with international standards and norms, and the risk environment scores among all 195 countries assessed are 40.2, 34.8, 41.9, 38.4, 26.4, 48.5, and 55.0 of a possible score of 100, respectively. All scores were normalized to a scale of 0 to 100, where 100 considered as the best health security conditions. Then according to the scores obtained by different countries, we classified them to highest (most prepared), middle (more prepared), and lowest (least prepared) countries.

### Data source

Data from global databases including Worldometer, WHO ^7^, Disease Control and Prevention Center (CDC) (16), and the Weekly Report on Complications of Death and Death (prepared by the CDC) were collected and retrieved according to the user guide of data sources for disease registration ^8^. Due to the rapid increase in data, the analysis in this study was conducted on April 14, 2020. According to the raw data of countries, the case fatality and recovery rates and their relationship is a unique feature of the GHS index, which provides a complete assessment of countries’ health security and a broader context of biological hazards in each country based on the geopolitical considerations of a country and its health system, as well as whether it has tested its capacity to control the spread of the disease. Thus, the case fatality and rate of recovery rates were calculated, then their relationship with GHS index and its six sub-component of GHS were investigated to provide a complete assessment in each country and its health system. Then, the country’s resilience to biological threats has been compared for countries with 1000 and above COVID-19 confirmed cases ^3^.

### Statistical analysis

Continuous variables are reported as range and mean ±SD. Categorical data are expressed as number (percentage). The univariate association between GHS index (and each of its components) with the number of total COVID-19 diagnostic tests, CFR and RR was assessed with correlation coefficient tests. Linear regression models were conducted to examine the association between the explanatory variables and the outcomes of interest. Ridge regression was applied to assess the impact of each parameter on total Covid-19 diagnostics tests controlling for the effects of population and GPD parameters (Model I). Multiple ridge regression analyses were used to determine parameters most predictive of the number of total COVID-19 diagnostic tests (Model II). Graphs were made in Excel 2013 and SPSS22 (IBM Corp. Released 2013. IBM SPSS Statistics for Windows, Version 22.0. Armonk, NY: IBM Corp). Correlation and regression analyses were performed using the statistical software Stata, Version 12 (StataCorp. 2011. *Stata Statistical Software: Release 12*. College Station, TX: StataCorp LP.). P-values less than 0.05 were considered statistically significant.

## Results

This study recruited data on 210 countries and territories, of which up to 14 April 2020, 72 countries (34.28%) with more than 1000 total COVID-19 cases were presents (**Appendix, Table S1**). Descriptive statistics of parameters is presented in **Table 1**.

**Table 1:** Characteristics of countries with more than 1000 total cases.

| Characteristics | Range | Mean ( ± SD) |
| --- | --- | --- |
| Population; million | 0.36 to 1386 | 80.71 (228.38) |
| GDP | 0.01 to 19.39 | 1.26 (3.05) |
| Total Cases | 1010 to 587155 | 26369.86 (76379.09) |
| Total Death | 2 to 23644 | 1652.38 (4630.98) |
| Total Recovered | 25 to 77738 | 6102.27 (15774.32) |
| Active Cases | 583 to 526563 | 18612.29 (64612.20) |
| Serious Critical | 1 to 12772 | 754.54 (2065.39) |
| Number of total COVID-19 diagnostic tests | 3359 to 2943955 | 204228.02 (425866.98) |
| CFR | 0.15 to 15.78 | 3.92 (3.50) |
| RR | 0.15 to 94.52 | 21.95 (19.58) |
| Prevention | 19.40 to 83.10 | 50.005 (14.34) |
| Detection | 12 to 98.20 | 60.03 (20.08) |
| Response | 19.50 to 91.90 | 50.69 (14.47) |
| Health System | 11 to 73 | 42.87 (14.40) |
| Capacity | 25.80 to 85.30 | 54.77 (13.85) |
| Risk Environment | 29.20 to 87.10 | 65.77 (12.85) |
| GHS Index | 23.60 to 83.50 | 53.37 (12.82) |

Abbreviations: GDP, Gross Domestic Production; CFR, Case Fatality Rate; RR, Recovery Rate; GHS, Global Health Security.

The frequency distribution of countries according to capacities to prevent, detect, response, health system, capacity, risk environment and general health score is presented in **figure 1**. In this figure level 1 represents the lowest and the level 5 representative of the highest capacity. Nineteen (26.38%) of 72 countries had overall health safety capacity at level 4 and 5 (**Figure** 1). Moreover, 14 (19.45%) countries had relatively high capacities to prevent, 36 (50%) had relatively high capacities to detect, 18 (25%) had relatively high capacities to respond, 10 (13.89%) had relatively high capacities in health system, 27 (37.5%) had relatively high compliance with international standards, and 41 (56.95%) had relatively high risk environment (**Figure 1**).

**Figure 1.**
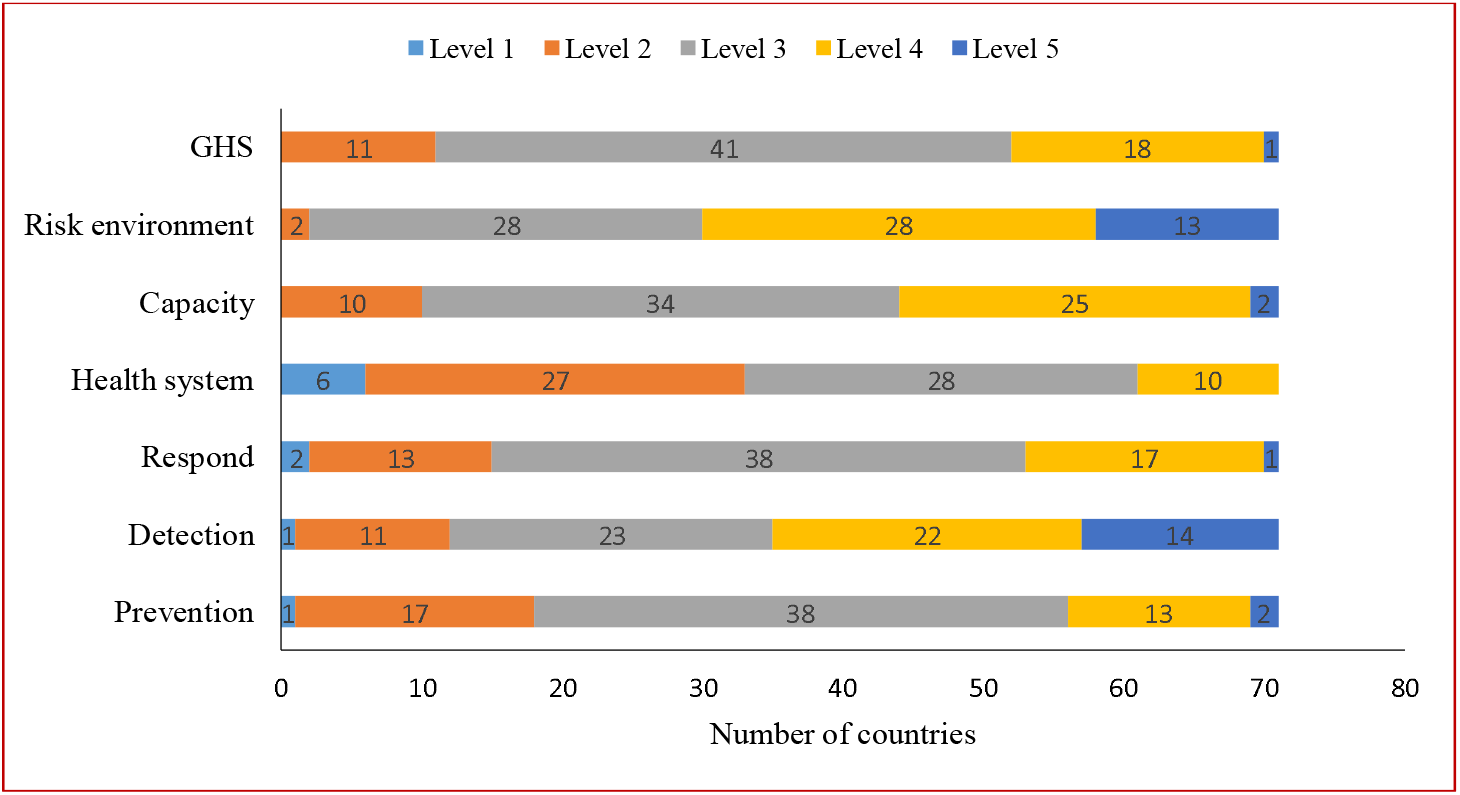
Number of studied countries according to capacities to prevent, detect, respond, health system, capacity, risk environment and general health score. Level 1 represents the lowest capacity and level 5 the highest.

The highest overall health safety capacity at level 4 and 5, were observed in Europe in thirteen (18.05%) countries (**Figure 2**).

**Figure 2.**
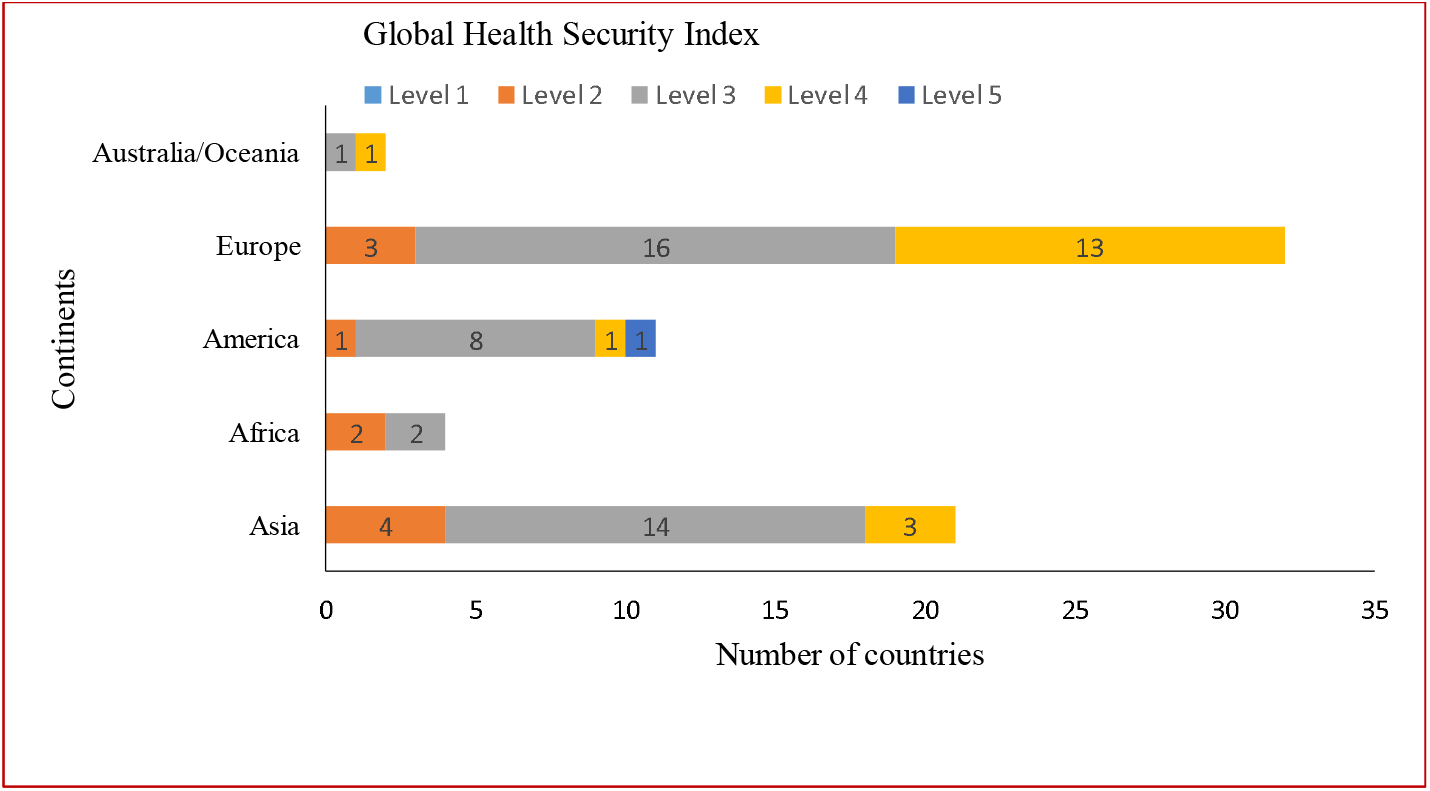
Global Health security Index by continents. Level 1 represents the lowest capacity and level 5 the highest.

The highest overall health safety capacity at level 4 and 5, were observed in high-income countries (**Figure 3**).

**Figure 3.**
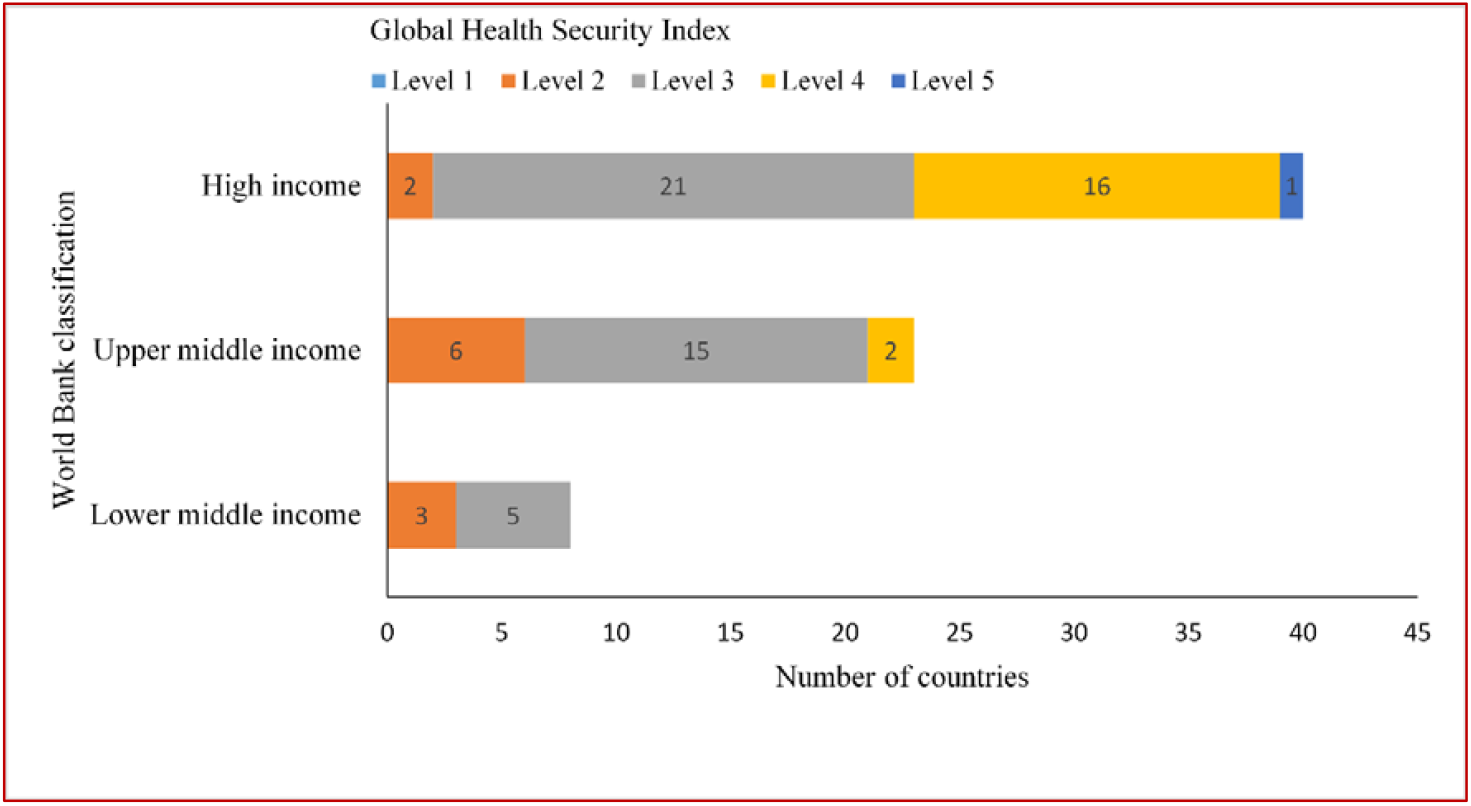
Global health security Index by World Bank classification. Level 1 represents the lowest capacity and level 5 the highest.

There was no significant association between the GHS index (and its components) with both of case fatality rate and recovery rate (**Appendix, Table S2**). The number of total COVID-19 diagnostic tests were positively correlated with GHS index (r=0.363; p=0.002) and all of its components except for the risk environment index (**Appendix, Table S2**).

According to the result of univariate linear regression models there was significant association between GHS index (**Figure 4)** and all of its components (**Figures 1-6, Appendix**) with number of total COVID-19 diagnostic tests (**Table S3, Appendix**).

**Figure 4.**
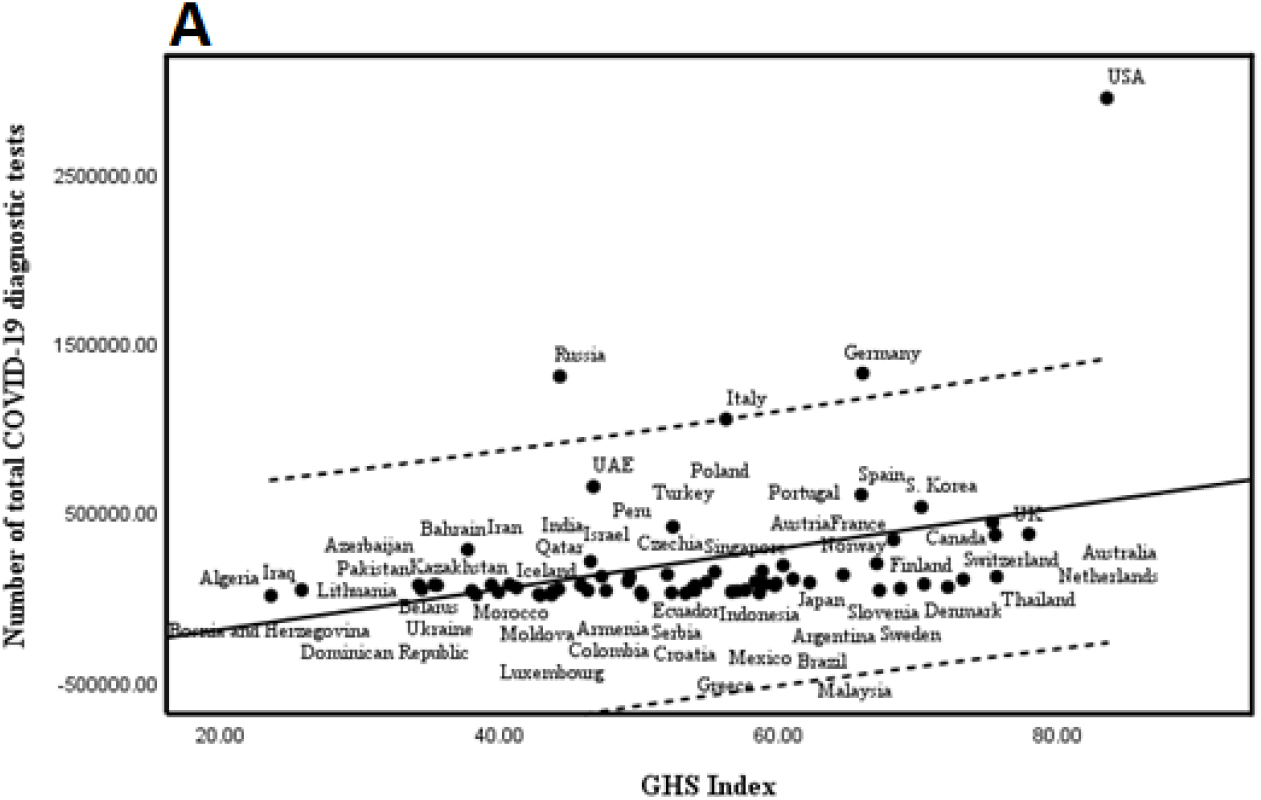
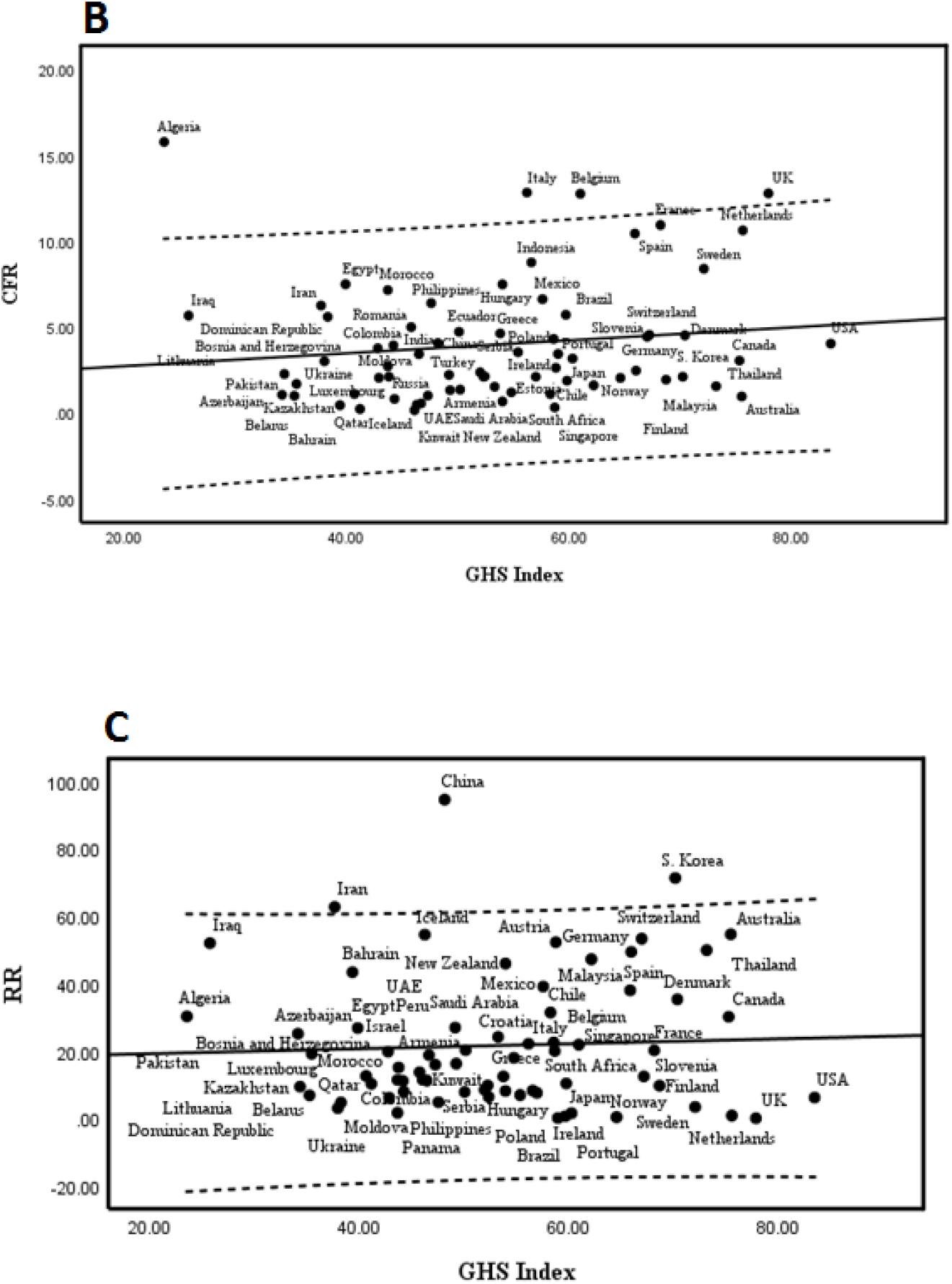
Scatter plot with regression line for GHS index and A, number of COVID-19 diagnostic tests, B, case fatality rate, and C, recovery rate

Using ridge regression, no significant association was found between the parameters and number of total COVID-19 diagnostic tests after controlling for the effects of population and GDP (**Table 2**).

**Table 2.**
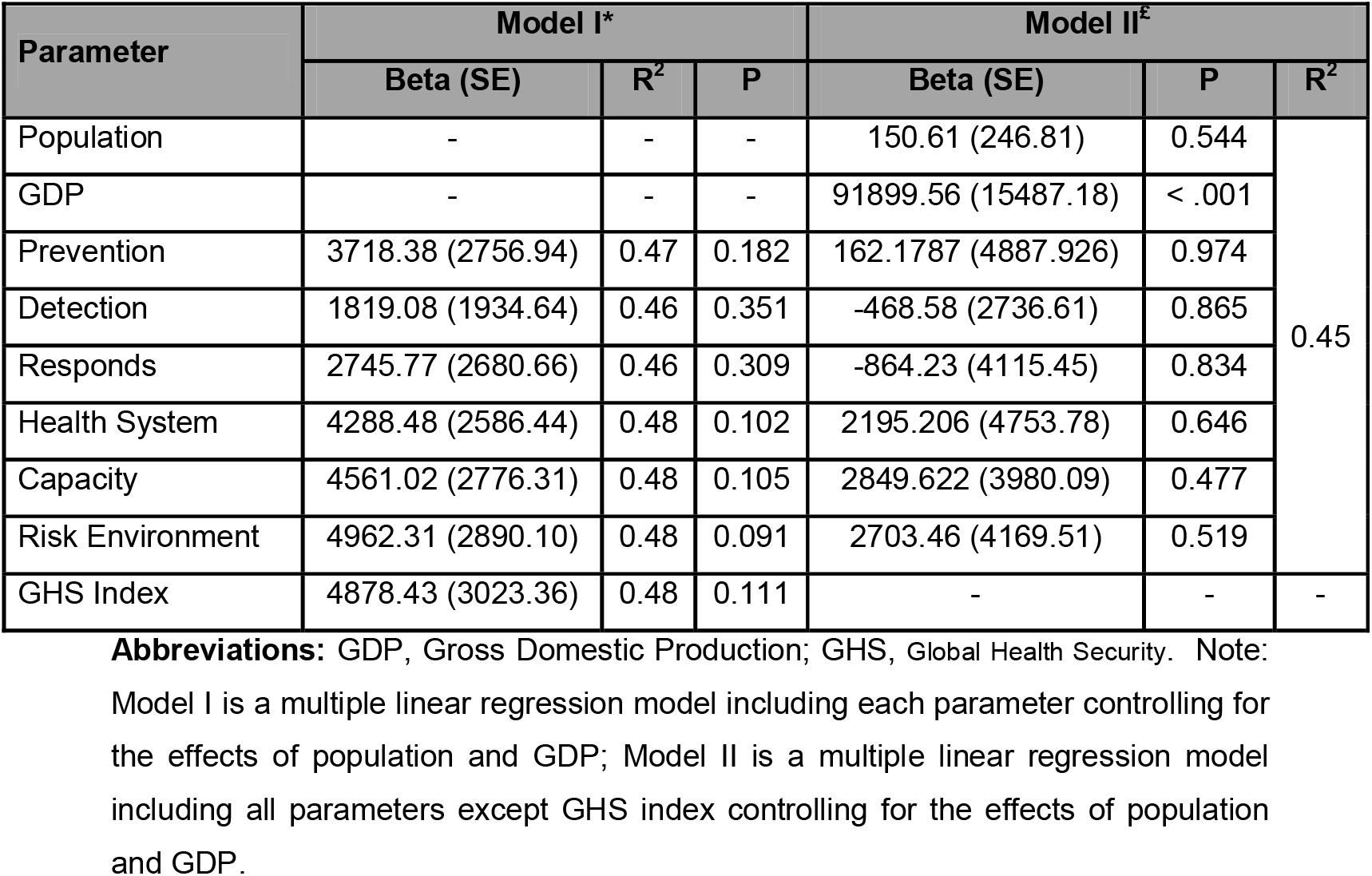
Results of multiple ridge regression analyses to determine parameters most predictive of the number of total COVID-19 diagnostic tests

Countries were divided in three categories according to their preparedness levels to face pandemics. In “most prepared group”, number of total COVID-19 diagnostic tests had a significant positive relation with GHS index (r=0.713; p=0.006). Recovery rate was negatively in relation to prevention index (r=-0.593; p=0.033). Case fatality rate was directly associated with the detection index (r=0.304; p=0.023) in “more prepared group” (**Table 3**).

**Table 3.** Association between GHS index and its component with the number of total COVID-19 diagnostic tests, CFR and RR in different preparedness levels

| Preparedness level |  | Number of total COVID-19 diagnostic tests |  |  | CFR |  |  | RR |  |  |
| --- | --- | --- | --- | --- | --- | --- | --- | --- | --- | --- |
|  | Parameter | CC | 95% CI for CC | P | CC | 95% CI for CC | P | CC | 95% CI for CC | P |
| Most prepared countries (n=13) | Prevention | 0.382 | (-0.214, 0.771) | 0.197 | 0.193 | (-0.400, 0.673) | 0.527 | -0.593 | (-0.862, -0.062) | 0.033 |
|  | Detection | 0.457 | (-0.125, 0.805) | 0.116 | -0.036 | (-0.576, 0.525) | 0.906 | 0.046 | (-0.518, 0.582) | 0.882 |
|  | Response | 0.281 | (-0.320, 0.720) | 0.353 | 0.311 | (-0.290, 0.736) | 0.301 | -0.148 | (-0.646, 0.439) | 0.629 |
|  | Health. System | 0.505 | (-0.064, 0.826) | 0.078 | -0.210 | (-0.682, 0.385) | 0.491 | 0.087 | (-0.487, 0.609) | 0.776 |
|  | Capacity | 0.545 | (-0.008, 0.843) | 0.054 | -0.210 | (-0.682, 0.386) | 0.491 | -0.286 | (-0.723, 0.315) | 0.344 |
|  | Risk. Environment | 0.015 | (-0.541, 0.561) | 0.962 | 0.285 | (-0.315, 0.723) | 0.345 | -0.258 | (-0.708, 0.342) | 0.395 |
|  | GHS Index | 0.713 | (0.268, 0.908) | 0.006 | 0.113 | (-0.467, 0.625) | 0.713 | -0.309 | (-0.735, 0.292) | 0.305 |
| More prepared countries (n=56) | Prevention | 0.212 | (-0.059, 0.454) | 0.123 | 0.150 | (-0.117, 0.397) | 0.269 | 0.035 | (-0.230, 0.295) | 0.799 |
|  | Detection | 0.096 | (-0.176, 0.355) | 0.488 | 0.304 | (0.044, 0.525) | 0.023 | 0.028 | (-0.237, 0.289) | 0.839 |
|  | Response | 0.172 | (-0.100, 0.421) | 0.212 | 0.106 | (-0.162, 0.359) | 0.439 | 0.018 | (-0.246, 0.279) | 0.896 |
|  | Health. System | 0.112 | (-0.161, 0.369) | 0.420 | 0.129 | (-0.138, 0.379) | 0.342 | 0.167 | (-0.101, 0.412) | 0.220 |
|  | Capacity | 0.193 | (-0.079, 0.438) | 0.162 | 0.199 | (-0.067, 0.439) | 0.141 | -0.142 | (-0.390, 0.126) | 0.298 |
|  | Risk. Environment | 0.143 | (-0.130, 0.395) | 0.303 | -0.060 | (-0.318, 0.206) | 0.658 | 0.193 | (-0.073, 0.434) | 0.154 |
|  | GHS Index | 0.221 | (-0.049, 0.462) | 0.108 | 0.245 | (-0.019, 0.477) | 0.068 | 0.084 | (-0.183, 0.339) | 0.539 |
| Least prepared countries (n=3) | --- | --- | --- | --- | --- | --- | --- | --- | --- | --- |

**Abbreviations:** GHS, Global Health Security; CC: Correlation Coefficient; CI: Confidence Interval; CFR: Case Fatality Rate; RR: Recovery Rate. The analyses were not performed in subgroups of “Least prepared countries” because of insufficient data.

According to different World Bank countries classifications, countries with more than 1000 total cases were divided in three categories. In “Lower-middle-income economies” group, case fatality rate positively related to detection, response and risk environment indices. Recovery rate has negative association with health system index (r = -0.637; p = 0.09). Case fatality rate negatively related to response, health system and GHS indices in “Upper-middle-income economies” group. In “High-income economies” group case fatality rate and number of total COVID-19 diagnostic tests were positively related to GHS index and its components except for risk environment index (**Table 4**).

**Table 4.** GHS index and its component association with the number of total COVID-19 diagnostic tests, CFR and RR in different World Bank countries classification

| World Bank countries classification |  | Number of total COVID-19 diagnostic tests |  |  | CFR |  |  | RR |  |  |
| --- | --- | --- | --- | --- | --- | --- | --- | --- | --- | --- |
|  | Parameters | CC | 95% CI for CC | P | CC | 95% CI for CC | P | CC | 95% CI for CC | P |
| Lower-middle-income economies (n=8) | Prevention | -0.325 | (-0.838, 0.492) | 0.432 | 0.401 | (-0.423, 0.862) | 0.325 | -0.491 | (-0.888, 0.327) | 0.217 |
|  | Detection | -0.129 | (-0.764, 0.633) | 0.760 | 0.716 | (0.024, 0.944) | 0.046 | -0.295 | (-0.827, 0.518) | 0.479 |
|  | Response | 0.355 | (-0.466, 0.848) | 0.388 | 0.714 | (0.019, 0.944) | 0.047 | 0.180 | (-0.601, 0.785) | 0.669 |
|  | Health. System | 0.369 | (-0.454, 0.852) | 0.369 | 0.096 | (-0.653, 0.750) | 0.821 | -0.637 | (-0.926, 0.123) | 0.090 |
|  | Capacity | -0.091 | (-0.748, 0.656) | 0.831 | 0.059 | (-0.673, 0.733) | 0.889 | -0.326 | (-0.838, 0.492) | 0.431 |
|  | Risk. Environment | 0.080 | (-0.662, 0.743) | 0.852 | 0.771 | (0.146, 0.956) | 0.025 | 0.230 | (-0.566, 0.804) | 0.583 |
|  | GHS Index | 0.061 | (-0.673, 0.734) | 0.886 | 0.637 | (-0.122, 0.926) | 0.089 | -0.371 | (-0.853, 0.452) | 0.366 |
| Upper-middle-income economies (n=23) | Prevention | 0.073 | (-0.360, 0.480) | 0.746 | -0.333 | (-0.655, 0.092) | 0.120 | 0.015 | (-0.400, 0.424) | 0.947 |
|  | Detection | -0.191 | (-0.567, 0.251) | 0.394 | -0.281 | (-0.621, 0.148) | 0.194 | 0.066 | (-0.356, 0.466) | 0.764 |
|  | Response | 0.078 | (-0.356, 0.483) | 0.731 | -0.432 | (-0.717, -0.025) | 0.039 | -0.048 | (-0.451, 0.371) | 0.827 |
|  | Health. System | 0.070 | (-0.362, 0.478) | 0.756 | -0.378 | (-0.683, 0.041) | 0.076 | 0.140 | (-0.289, 0.522) | 0.525 |
|  | Capacity | 0.089 | (-0.345, 0.492) | 0.692 | -0.301 | (-0.635, 0.126) | 0.162 | -0.090 | (-0.484, 0.335) | 0.683 |
|  | Risk. Environment | -0.093 | (-0.496, 0.342) | 0.679 | -0.214 | (-0.576, 0.217) | 0.326 | -0.023 | (-0.431, 0.393) | 0.919 |
|  | GHS Index | -0.009 | (-0.429, 0.414) | 0.967 | -0.413 | (-0.705, -0.001) | 0.050 | 0.021 | (-0.394, 0.430) | 0.923 |
| High-income economies (n=41) | Prevention | 0.364 | (0.054, 0.609) | 0.023 | 0.425 | (0.131, 0.650) | 0.006 | -0.070 | (-0.373, 0.247) | 0.669 |
|  | Detection | 0.382 | (0.075, 0.622) | 0.016 | 0.391 | (0.090, 0.626) | 0.013 | 0.093 | (-0.225, 0.393) | 0.569 |
|  | Response | 0.338 | (0.025, 0.591) | 0.035 | 0.365 | (0.060, 0.607) | 0.021 | 0.043 | (-0.272, 0.350) | 0.793 |
|  | Health. System | 0.326 | (0.012, 0.582) | 0.043 | 0.387 | (0.086, 0.623) | 0.014 | 0.140 | (-0.179, 0.433) | 0.388 |
|  | Capacity | 0.410 | (0.108, 0.642) | 0.010 | 0.338 | (0.030, 0.588) | 0.033 | -0.086 | (-0.387, 0.232) | 0.598 |
|  | Risk. Environment | 0.115 | (-0.208, 0.416) | 0.485 | 0.165 | (-0.154, 0.453) | 0.309 | 0.196 | (-0.123, 0.478) | 0.226 |
|  | GHS Index | 0.427 | (0.128, 0.654) | 0.007 | 0.447 | (0.157, 0.666) | 0.004 | 0.083 | (-0.234, 0.385) | 0.610 |

**Abbreviations:** GHS, Global Health Security; CFR: Case Fatality Rate; RR: Recovery Rate; CC: Correlation Coefficient; CI: Confidence Interval.

According to different cut off for GHS index and its components, studied countries were divided. Bivariate correlation between GHS index and outcomes of interest in each subgroups is shown in **table S6** (**Appendix**). Table S7 represented association between each index with outcomes in subgroups according to GHS cut off (**Appendix**).

Countries were divided in five categories according to their continent. In Asia, case fatality rate had a significant negative relation with risk environment index (r=-0.489; p=0.021). Recovery rate was positively in relation to risk environment index, in European countries (r=0.353; p=0.048). Also, case fatality rate was directly associated with the GHS index (r=0.547; p=0.001) and its component except for risk environment index. There was a significant association between the GHS index (and its components except for detection index) with the number of total COVID-19 diagnostic tests in America (**Table 5**).

**Table 5.**
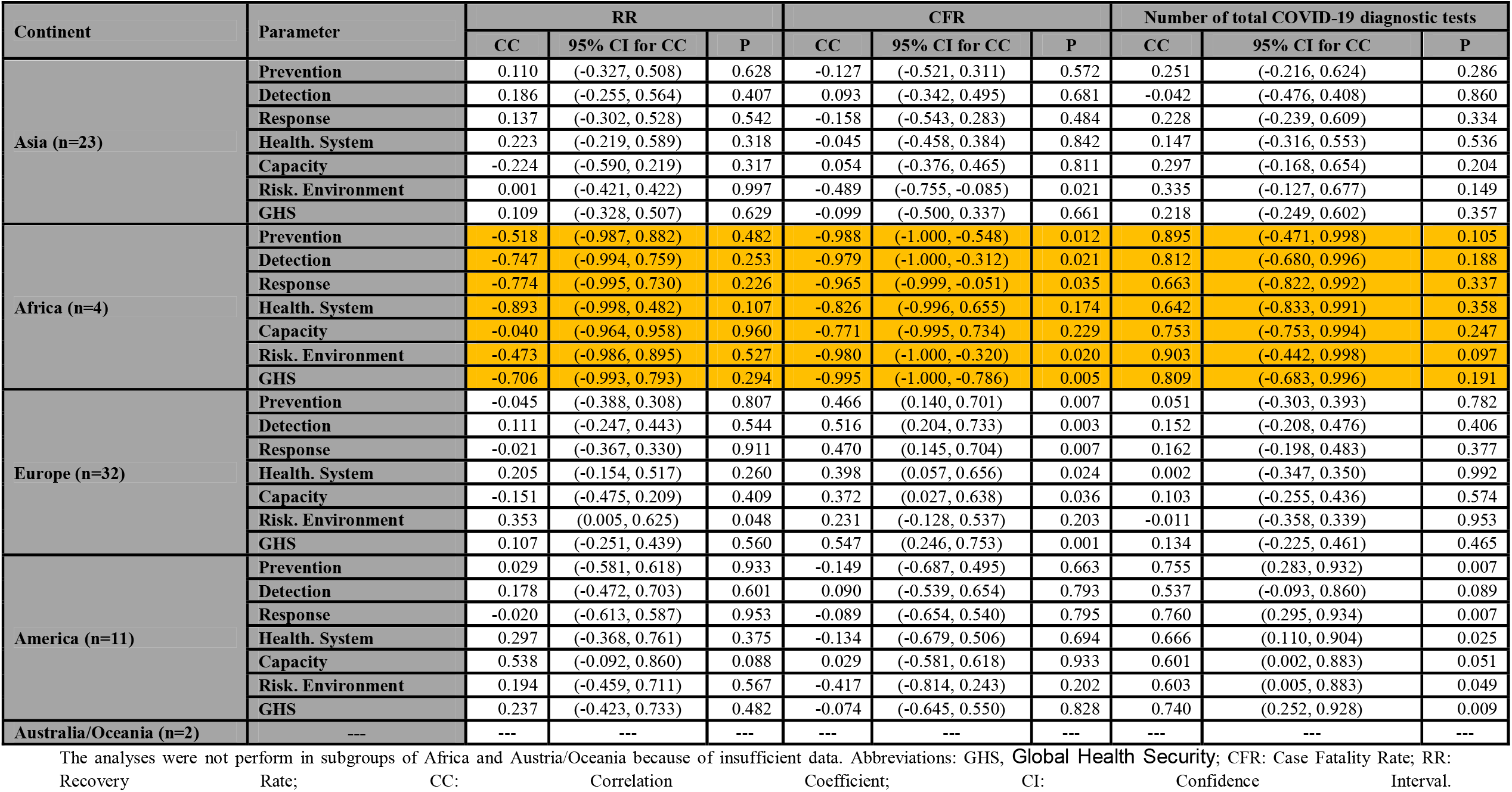
Association between GHS index and its component with the number of total COVID-19 diagnostic tests, CFR and RR in different Continents

| Continent | Parameter | RR |  |  | CFR |  |  | Number of total COVID-19 diagnostic tests |  |  |
| --- | --- | --- | --- | --- | --- | --- | --- | --- | --- | --- |
|  |  | CC | 95% CI for CC | P | CC | 95% CI for CC | P | CC | 95% CI for CC | P |
| Asia (n=23) | Prevention | 0.110 | (-0.327, 0.508) | 0.628 | -0.127 | (-0.521, 0.311) | 0.572 | 0.251 | (-0.216, 0.624) | 0.286 |
|  | Detection | 0.186 | (-0.255, 0.564) | 0.407 | 0.093 | (-0.342, 0.495) | 0.681 | -0.042 | (-0.476, 0.408) | 0.860 |
|  | Response | 0.137 | (-0.302, 0.528) | 0.542 | -0.158 | (-0.543, 0.283) | 0.484 | 0.228 | (-0.239, 0.609) | 0.334 |
|  | Health. System | 0.223 | (-0.219, 0.589) | 0.318 | -0.045 | (-0.458, 0.384) | 0.842 | 0.147 | (-0.316, 0.553) | 0.536 |
|  | Capacity | -0.224 | (-0.590, 0.219) | 0.317 | 0.054 | (-0.376, 0.465) | 0.811 | 0.297 | (-0.168, 0.654) | 0.204 |
|  | Risk. Environment | 0.001 | (-0.421, 0.422) | 0.997 | -0.489 | (-0.755, -0.085) | 0.021 | 0.335 | (-0.127, 0.677) | 0.149 |
|  | GHS | 0.109 | (-0.328, 0.507) | 0.629 | -0.099 | (-0.500, 0.337) | 0.661 | 0.218 | (-0.249, 0.602) | 0.357 |
| Africa (n=4) | Prevention | -0.518 | (-0.987, 0.882) | 0.482 | -0.988 | (-1.000, -0.548) | 0.012 | 0.895 | (-0.471, 0.998) | 0.105 |
|  | Detection | -0.747 | (-0.994, 0.759) | 0.253 | -0.979 | (-1.000, -0.312) | 0.021 | 0.812 | (-0.680, 0.996) | 0.188 |
|  | Response | -0.774 | (-0.995, 0.730) | 0.226 | -0.965 | (-0.999, -0.051) | 0.035 | 0.663 | (-0.822, 0.992) | 0.337 |
|  | Health. System | -0.893 | (-0.998, 0.482) | 0.107 | -0.826 | (-0.996, 0.655) | 0.174 | 0.642 | (-0.833, 0.991) | 0.358 |
|  | Capacity | -0.040 | (-0.964, 0.958) | 0.960 | -0.771 | (-0.995, 0.734) | 0.229 | 0.753 | (-0.753, 0.994) | 0.247 |
|  | Risk. Environment | -0.473 | (-0.986, 0.895) | 0.527 | -0.980 | (-1.000, -0.320) | 0.020 | 0.903 | (-0.442, 0.998) | 0.097 |
|  | GHS | -0.706 | (-0.993, 0.793) | 0.294 | -0.995 | (-1.000, -0.786) | 0.005 | 0.809 | (-0.683, 0.996) | 0.191 |
| Europe (n=32) | Prevention | -0.045 | (-0.388, 0.308) | 0.807 | 0.466 | (0.140, 0.701) | 0.007 | 0.051 | (-0.303, 0.393) | 0.782 |
|  | Detection | 0.111 | (-0.247, 0.443) | 0.544 | 0.516 | (0.204, 0.733) | 0.003 | 0.152 | (-0.208, 0.476) | 0.406 |
|  | Response | -0.021 | (-0.367, 0.330) | 0.911 | 0.470 | (0.145, 0.704) | 0.007 | 0.162 | (-0.198, 0.483) | 0.377 |
|  | Health. System | 0.205 | (-0.154, 0.517) | 0.260 | 0.398 | (0.057, 0.656) | 0.024 | 0.002 | (-0.347, 0.350) | 0.992 |
|  | Capacity | -0.151 | (-0.475, 0.209) | 0.409 | 0.372 | (0.027, 0.638) | 0.036 | 0.103 | (-0.255, 0.436) | 0.574 |
|  | Risk. Environment | 0.353 | (0.005, 0.625) | 0.048 | 0.231 | (-0.128, 0.537) | 0.203 | -0.011 | (-0.358, 0.339) | 0.953 |
|  | GHS | 0.107 | (-0.251, 0.439) | 0.560 | 0.547 | (0.246, 0.753) | 0.001 | 0.134 | (-0.225, 0.461) | 0.465 |
| America (n=11) | Prevention | 0.029 | (-0.581, 0.618) | 0.933 | -0.149 | (-0.687, 0.495) | 0.663 | 0.755 | (0.283, 0.932) | 0.007 |
|  | Detection | 0.178 | (-0.472, 0.703) | 0.601 | 0.090 | (-0.539, 0.654) | 0.793 | 0.537 | (-0.093, 0.860) | 0.089 |
|  | Response | -0.020 | (-0.613, 0.587) | 0.953 | -0.089 | (-0.654, 0.540) | 0.795 | 0.760 | (0.295, 0.934) | 0.007 |
|  | Health. System | 0.297 | (-0.368, 0.761) | 0.375 | -0.134 | (-0.679, 0.506) | 0.694 | 0.666 | (0.110, 0.904) | 0.025 |
|  | Capacity | 0.538 | (-0.092, 0.860) | 0.088 | 0.029 | (-0.581, 0.618) | 0.933 | 0.601 | (0.002, 0.883) | 0.051 |
|  | Risk. Environment | 0.194 | (-0.459, 0.711) | 0.567 | -0.417 | (-0.814, 0.243) | 0.202 | 0.603 | (0.005, 0.883) | 0.049 |
|  | GHS | 0.237 | (-0.423, 0.733) | 0.482 | -0.074 | (-0.645, 0.550) | 0.828 | 0.740 | (0.252, 0.928) | 0.009 |
| Australia/Oceania (n=2) | --- | --- | --- | --- | --- | --- | --- | --- | --- | --- |

The analyses were not perform in subgroups of Africa and Austria/Oceania because of insufficient data. Abbreviations: GHS, Global Health Security; CFR: Case Fatality Rate; RR: Recovery Rate; CC: Correlation Coefficient; CI: Confidence Interval.

## Discussion

This study aimed to evaluate the GHS index in countries affected by the COVID-19 outbreak and its public health related consequences, as well as its relationship with the case fatality and recovery rates, showed a wide variety between countries in term of prevention, detection and response to COVID-19 disease. In line with the present study, recently a published report of the International Health Regulations (IHR) on data from 182 countries preparedness, capacity building and collaboration between countries are still need improvement, as well as local readiness for outbreak control needed to strengthen ^1^. Numerous factors affect the COVID-19 disease emergence and spread within countries and between geographical regions, comprising the national capacities, the ability of detection, prevention and control, climate-related factors, and population’s density ^9-11^. One of the important factors in the context of GHS index is health system, our results revealed a significant correlation between the number of COVID-19 tests and health security capacities ^12,13^. Other aspects of such important variable is implementing public health prevention strategies, such as public awareness about hand hygiene and social distancing ^14,15^. Our analysis shed the light on the fact that most countries at the top of the list with the highest number of cases, as well as deaths from COVID-19, were in more prepared condition according to the GHS index. This may be point to this issue that health security is essentially weak in the national as well as global level ^16^; thus, no country or region was fully prepared ho handle COVID-19 epidemics and pandemics, and each country and region has its own gaps to fill. Many countries have been ranked as region with low capacities to detect, prevent and combat the outbreak, through their experience with former infectious diseases ^17,18^. This fact may indicate more than ever that a special attention on regions of low human and health development are needed, as well as further improvements in observation with additional international collaboration are required ^19,20^.

The WHO claimed that one of the reasons for the large difference of mortality rate among various countries, could be the difference in life expectancy, medical facilities, and the number of tests performed. Though Europe and America has become the center of the COVID-19 outbreak in the world now, some countries in these regions such as Belarus is the only country on the European continent whose officials are not seeking fundamental changes in people’s daily lives. These country has taken a very different approach to the combat COVID-19 than other European countries and even its closest neighbors such as Russia and Ukraine. While Ukraine is close to declaring a state of emergency, and Russia has closed public places universities and schools, as well as canceled public events, and stopped all incoming and outgoing flights ^21^. Interestingly, the highest prepared region according to the GHS index was Europe with the most frequent level 4 and above results. Although European countries account for the highest number of COVID-19 confirmed cases and related deaths, this can be attributed in part to the high number of hospital and intensive care unit (ICU) beds ^22^. Besides, these countries also quickly conducted reliable screening and diagnostic tests for the COVID-19 disease and implemented strict social distancing measures to prevent the outbreak ^23^. Another possible reason of the rapid test for COVID-19 could be that in some of these countries, such as Germany, private laboratories across the country were free to provide testing and have helped the country test millions of people for the disease.

The most important point is that the condition of health-care system, Hospital and ICU beds and mechanical ventilation facilities of a country and its preparedness, which can slow-down the course of the COVID-19 disease outbreak, is in what situation; therefore, a sufficient, well-equipped and well-prepared health-care system in a country is crucial for severe patients infected by COVID-19 disease. Lessons learned from the previous infectious disease spread and emergencies, such as SARS-CoV, MERS-CoV, H1N1 flu, Zika, and Ebola viruses’ outbreak, have lead several countries to make substantial improvement in effective detection, prevention, and the ability to response to similar conditions ^24-26^. One of the key component that lead to increasing the ability of a country to response quickly to the disease outbreak is early detection by the development of laboratories equipped specifically for COVID-19 disease. We observed 50% of countries with robust detection capacities, mostly felled in the European and Asian continents, as well as were from high-income countries. The rapid response capacity rely on the level of preparedness of a country in term of availability of health system resources and emergency logistics. The present study findings showed that most of countries need more support by national as well as global actions to more strengthen health security. Besides, it seems that many countries, especially low-income countries, not only has not increased their capacity and readiness, but also underline the importance of the investment in the ability to combat the disease outbreak ^27^.

COVID-19 testing provides two functions, including to diagnose people infected with the virus, and monitoring and surveillance, especially in cases where the symptoms are mild or not obvious at all. We observed a significant association between numbers of COVID-19 testes performed and case fatality rate, as well as GHS index. This means that in most prepared countries to deal with the disease outbreak, more will be tested in less time and as a result more will be diagnosed ^28^. Thus, less testing or insufficient testing at the proper time may decelerating resourceful patient care and sending a heavily lagged view of the COVID-19 outbreak to health-care system and its decision makers.

## Limitations

Even with clear and precise descriptions for each of GHS index components and the index itself, finding the relevant data points remains controversial. Thus, while quicker detection and response to COVID-19 outbreaks help to diminish the total number of cases, it is still unclear whether process improvements, which may be reflected in measuring GHS index, will improve health outcomes in a population.

## Conclusion

Measuring the GHS index for each country as well as globally, may help to create progress in faster detection and response to a disease outbreaks, as well as improve the health security at national and global levels. But it is necessary to work with countries and their regional networks to improve the approach to share accurate and up-to-date local knowledge with the global health community. In light of necessities to enhance local as well as global capacities, preparedness and response to the COVID-19 outbreak, future efforts to measure global health security might be beneficial when using complementary modes of data collection. This might also be useful in further refining those capacities and capabilities that are necessary in countries with poor healthcare and public health infrastructure. Finally, this shows the importance of timely and comprehensive evaluations and emphasizes that even more must be done to build surge capacity to prevent, detect, and respond to such emergencies around the world.

## Data Availability

all data presented in the manuscript.

## Authors’ Contribution

E.M and F.R developed the original idea and the protocol, abstracted, analyzed the data, wrote the manuscript, and were guarantors. F.R and A.S.M developed the protocol, abstracted the data, and prepared the manuscript.

## Ethical Approval

We performed a secondary analysis of the data without Institutional Review Board approval.

## References

1. Kandel N, Chungong S, Omaar A, Xing J. Health security capacities in the context of COVID-19 outbreak: an analysis of International Health Regulations annual report data from 182 countries. Lancet (London, England). 2020;395(10229):1047–1053.

2. Vellingiri B, Jayaramayya K, Iyer M, et al. COVID-19: A promising cure for the global panic. The Science of the total environment. 2020;725:138277.

3. Worlometers. https://www.worldometers.info/coronavirus/.Published 2020. Accessed 10 March 2020.

4. Yi Y, Lagniton PNP, Ye S, Li E, Xu RH. COVID-19: what has been learned and to be learned about the novel coronavirus disease. International journal of biological sciences. 2020;16(10):1753–1766.

5. Guan W-j, Ni Z-y, Hu Y, et al. Clinical Characteristics of Coronavirus Disease 2019 in China. 2020.

6. Yang XH, Sun RH, Chen DC. [Diagnosis and treatment of COVID-19: acute kidney injury cannot be ignored]. Zhonghua yi xue za zhi. 2020;100(0):E017.

7. World Health Organization (WHO). Coronavirus disease 2019 (COVID-19) Situation report – 43. Web site. Search ResultsWeb resultsCoronavirus disease 2019 (COVID-19) - World Health …www.who.int › docs › situation-reports › 20200303-sitrep-43-covid-19. Published 2020. Accessed.

8. Centers of Disease Control and Prevention (CDC). https://www.cdc.gov/coronavirus/2019-ncov/cases-in-us.html. Published 2020. Accessed March 10, 2020.

9. Geographic Differences in COVID-19 Cases, Deaths, and Incidence - United States, February 12-April 7, 2020. MMWR Morbidity and mortality weekly report. 2020;69(15):465–471.

10. Booth M. Climate Change and the Neglected Tropical Diseases. Adv Parasitol. 2018;100:39–126.

11. Smiley Evans T, Shi Z, Boots M, et al. Synergistic China–US Ecological Research is Essential for Global Emerging Infectious Disease Preparedness. EcoHealth. 2020;17(1):160–173.

12. Quinn SC, Kumar S. Health inequalities and infectious disease epidemics: a challenge for global health security. Biosecur Bioterror. 2014;12(5):263–273.

13. Hunsperger E, Juma B, Onyango C, et al. Building laboratory capacity to detect and characterize pathogens of public and global health security concern in Kenya. BMC Public Health. 2019;19(3):477.

14. Arbogast JW, Moore-Schiltz L, Jarvis WR, Harpster-Hagen A, Hughes J, Parker A. Impact of a Comprehensive Workplace Hand Hygiene Program on Employer Health Care Insurance Claims and Costs, Absenteeism, and Employee Perceptions and Practices. 2016;58(6):e231–e240.

15. Branch-Elliman W, Price CS, Bessesen MT, Perl TM. Using the Pillars of Infection Prevention to Build an Effective Program for Reducing the Transmission of Emerging and Reemerging Infections. Current Environmental Health Reports. 2015;2(3):226–235.

16. Heymann DL, Chen L, Takemi K, et al. Global health security: the wider lessons from the west African Ebola virus disease epidemic. Lancet (London, England). 2015;385(9980):1884–1901.

17. Benecke O, DeYoung SE. Anti-Vaccine Decision-Making and Measles Resurgence in the United States. Global pediatric health. 2019;6:2333794x19862949.

18. Tam T. Fifteen years post-SARS: Key milestones in Canada’s public health emergency response. Canada communicable disease report = Releve des maladies transmissibles au Canada. 2018;44(5):98–101.

19. Kluberg SA, Mekaru SR, McIver DJ, et al. Global Capacity for Emerging Infectious Disease Detection, 1996-2014. Emerg Infect Dis. 2016;22(10):E1–E6.

20. Gilbert M, Pullano G, Pinotti F, et al. Preparedness and vulnerability of African countries against importations of COVID-19: a modelling study. Lancet (London, England). 2020;395(10227):871–877.

21. Bedford J, Enria D, Giesecke J, et al. COVID-19: towards controlling of a pandemic. The Lancet. 2020;395(10229):1015–1018.

22. Grasselli G, Pesenti A, Cecconi M. Critical Care Utilization for the COVID-19 Outbreak in Lombardy, Italy: Early Experience and Forecast During an Emergency Response. JAMA. 2020.

23. Remuzzi A, Remuzzi G. COVID-19 and Italy: what next? The Lancet. 2020;395(10231):1225–1228.

24. Institute of Medicine Forum on Microbial T. The National Academies Collection: Reports funded by National Institutes of Health. In: Knobler S, Mahmoud A, Lemon S, Mack A, Sivitz L, Oberholtzer K, eds. Learning from SARS: Preparing for the Next Disease Outbreak: Workshop Summary. Washington (DC): National Academies Press (US) Copyright © 2004, National Academy of Sciences.; 2004.

25. Engman ML. SARS outbreak and lessons learned. Journal of insurance medicine (New York, NY). 2002;34(2):83–85.

26. Piot P, Soka MJ, Spencer J. Emergent threats: lessons learnt from Ebola. International Health. 2019;11(5):334–337.

27. WHO. Thematic paper on the status of country preparedness capacities. World Health Organization. http://apps.who.int/gpmb/assets/thematic_papers/tr-2.pdf Published 2019. Accessed 21 April, 2020.

28. Adepoju P. Nigeria responds to COVID-19; first case detected in sub-Saharan Africa. Nat Med. 2020;26(4):444–448.

